# Pharmacokinetics of remdesivir in a COVID-19 patient with end-stage renal disease on intermittent hemodialysis

**DOI:** 10.1101/2020.10.28.20216887

**Authors:** Fritz Sörgel, Jakob J. Malin, Henning Hagmann, Martina Kinzig, Muhammad Bilal, Dennis A. Eichenauer, Oliver Scherf-Clavel, Alexander Simonis, Lobna El Tabei, Uwe Fuhr, Jan Rybniker

## Abstract

Remdesivir, a drug with provisional approval for the treatment of COVID-19, is not recommended in patients with an estimated glomerular filtration rate ≤ 30 mL/min. Here we provide a first detailed pharmacokinetic assessment of remdesivir and its major metabolites in a patient with end stage renal disease on hemodialysis.

## Introduction

Remdesivir has been authorized for emergency use in patients with severe SARS-CoV-2 infection [1] as it reduced the median time to recovery from COVID-19 in a randomized controlled trial [2]. Due to a lack of data, remdesivir is not recommended for patients with an estimated glomerular filtration rate (eGFR) ≤ 30 mL/min, including those with end-stage renal disease (ESRD), which represents a substantial limitation for COVID-19 treatment in this vulnerable population. Patients with ESRD have an increased risk for a severe course of COVID-19 [3]. Moreover, COVID-19 itself leads to AKI and AKI-associated mortality in a significant proportion of critically ill patients [4]. Thus, it is of high importance to assess safety and pharmacokinetics of remdesivir in patients with renal impairment and those on renal replacement therapy [5].

Intracellular remdesivir prodrug (GS-5734) is rapidly converted to its alanine metabolite GS-704277 and subsequently to the monophosphate that is finally converted to the active triphosphate GS-443902 [6]. Alternatively, dephosphorylation of the monophosphate yields the remdesivir nucleoside core (GS-441524) which becomes the predominant circulating metabolite providing a potential source of active drug by a slow re-phosphorylation process [7]. In healthy volunteers, renal excretion of a remdesivir dose is about 10% as unchanged drug [8] and about 50 % as GS-441524 [9].

Here we report on pharmacokinetics of remdesivir and its metabolites and treatment outcome in a patient on renal replacement therapy without residual renal function suffering from severe COVID-19.

## Methods

### Administration of remdesivir

Remdesivir was administered as controlled intravenous infusion (500 mL/h) over 30 minutes via femoral vein catheter without any parallel infusions (for doses and timing see figure 1). Markers for hepatotoxicity were monitored at least daily during treatment with remdesivir and for subsequent days (supplementary figure 2).

**Figure 1.**
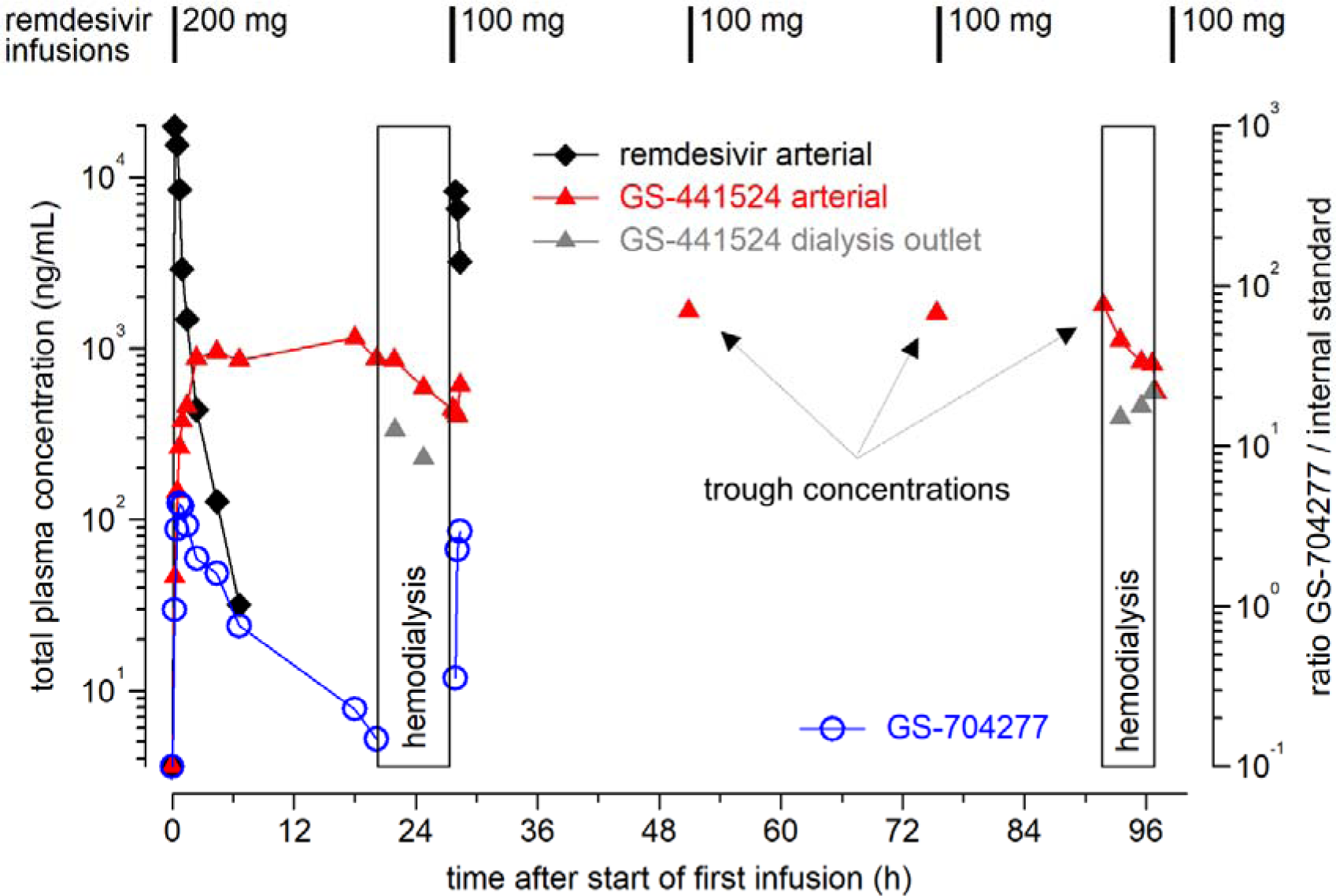
Pharmacokinetics of the prodrug remdesivir (GS-5734), its intermediate alanine metabolite (GS-704277) and the predominant circulating metabolite GS-441524 during a standard 5-day regimen in a patient with intermittent hemodialysis due to long lasting ESRD. Arterial blood samples were drawn in conjunction to initial administration of remdesivir and hemodialysis (d1, d4). In addition, GS-441524 concentrations in samples from dialysis outlet (gray triangles) are shown that were taken to assess elimination by hemodialysis.

### Pharmacokinetic sampling

Blood samples were drawn from the arterial line after discarding dead space volume. In addition, several blood samples were taken from the inlet and the outlet of the dialysis device to analyze drug elimination during hemodialysis. Polypropylene tubes containing 1.2 mg/mL ethylenediaminetetraacetic acid (EDTA) to prevent clotting and 1 mg/mL of sodium fluoride (Sarstedt, Nümbrecht, Germany) were used and directly placed on ice to prevent *in vitro* hydrolysis of remdesivir [10] and its metabolites before further processing within a maximum of one hour. Plasma was separated by centrifugation at 4°C and subsequently stored at −80°C prior to pharmacokinetic analyses.

### Hemodialysis

Hemodialysis was performed using the Fresenius GENIUS^®^ device, a single pass close tank hemodialysis system, containing 90 liters of dialysis fluid. The system was equipped with an FX600 CorDiax^®^ hemodialysis filter (Fresenius) and standard blood lines and tubing. The extracorporeal circuit was established via central venous catheter. Blood flow ranged between 150 and 280 mL/min. According to technical design, dialysate flow equaled blood flow. Ultrafiltration settings ranged between 100 and 200 ml/h. Hemodialysis was performed prior to administration of the daily remdesivir dose on days 2 and 5 of treatment. In the two dialysis sessions a blood volume of 88 and 90 liters was processed. Duration of treatment was 425 minutes and 315 minutes. Ultrafiltration volume was 990 ml and 1037 ml, respectively. For anticoagulation unfractionated heparin was administered as intravenous bolus of 2000 IE at the beginning of hemodialysis and a rate of 1000 IE/h during treatment.

### Determination of remdesivir and metabolites

Measurements of drug concentrations were performed at the Institute for Biomedical and Pharmaceutical Research, Nürnberg-Heroldsberg, Germany. Remdesivir and its metabolite GS-441524 were purchased from BIOSYNTH^®^ Carbosynth (Berkshire, UK). Calibration standards and spiked quality control samples were prepared in drug-free human EDTA plasma. Liquid chromatography/mass spectrometry to detect the compounds used transitions REM 603.3→402.0 and 292.2→163.1 (GS-441524) on a SCIEX API 6500TM triple quadrupole mass spectrometer equipped with turbo ion spray interface (SCIEX, Concord, Ontario, Canada). Calibration for both compounds was performed by weighted (1/concentration^2^) linear regression. Linearity for remdesivir and GS-441524 in human plasma could be demonstrated over a calibration range from 3.600 to 787.4 ng/mL for remdesivir and from 10.84 to 474.5 ng/mL for GS-441524. Quantification of remdesivir and GS-441524 was performed by peak area ratio of analyte to internal standard (remdesivir: piperacillin-d5, GS-441524 amoxicillin-d4). When samples were outside that range a dilution by drug free human plasma was used. No interferences were observed for remdesivir and GS-441524 and the internal standards in human plasma. Since there was no reference standard available for the intermediate metabolite GS-704277, only the time course of plasma concentrations could be described which were obtained from peak heights relative to an internal standard (transition 443.1→202.1).

### Pharmacokinetic assessment

Pharmacokinetic parameters following the first dose for remdesivir and its metabolites were calculated by standard non-compartmental methods. Extraction of GS-441524 by dialysis was calculated as (1-outlet concentration [corrected for volume loss] / inlet concentration). The observed decrease of GS-441524 extraction during dialysis was described by linear regression with cumulative blood flow.

## Results and Discussion

In September 2020, a male patient in his seventies was admitted to the Hospital and diagnosed with COVID-19 (Supplementary tables 1 and 2). Initially, he presented with typical pulmonary infiltrates and normal arterial blood gas results. However, clinical status deteriorated rapidly and dexamethasone treatment was initiated. At this time-point, remdesivir was not considered due to chronic renal impairment requiring renal replacement therapy. Sequential CT scans revealed progression of pulmonary infiltrates (Supplementary figure 1). Nine days after initial admission, the patient was transferred to the ICU with hypoxemic lung failure requiring high-flow oxygen therapy. In this critical situation, we decided to apply remdesivir while performing extensive therapeutic drug monitoring of the prodrug and two metabolites for safety reasons. Remdesivir concentrations decreased rapidly after the first infusion with an apparent half-life of 1.1 hours and were below the lower limit of quantification prior to dialysis (fig 1; supplementary tables 3 and 4). GS-441524 concentrations increased up to 1 µg/mL and remained essentially constant. Exposure for remdesivir (AUC_0-∞_ 13.0 µg/mL* h, C_max_ 19.8 µg/mL) and GS-441524 (AUC_0-20.1 h_ 18.4 µg/mL* h, C_max_ 1.15 µg/mL) after the first dose was about 3-fold and 6-fold higher in our patient as compared to healthy volunteers (Supplementary tables 3 and 4) [8] which is in agreement with a preliminary observation in a critically ill patient with reduced renal function [11]. Both remdesivir clearance and volume of distribution were clearly lower in our patient, which may be attributable to his low body weight (53.2 kg). Hemodialysis performed prior to the second and the 5^th^ dose of remdesivir reduced GS-441524 plasma concentrations by about 50% (fig. 1). The initial extraction rate of dialysis was 72% and decreased to 42% at the end of dialysis (Supplementary table 5). Trough concentrations of GS-441524 were high but stable between dialysis sessions (range 1.66 to 1.79 µg/mL), while remdesivir trough concentrations were always below the lower limit of quantification (fig. 1, supplementary tables 3 and 4). Thus, we did not observe significant accumulation of remdesivir and accumulation of GS-441524 was prevented by intermittent hemodialysis.

Accumulation of the vehicle sulfobutylether-beta-cyclodextrin (SBECD) may occur in renal failure [12] and high doses were associated with significant hepatic and renal toxicity in animals [13]. However, reported toxic doses were 50-100 times higher than exposure during a 5-10 day course of remdesivir. In addition, it was shown that SBECD is eliminated by renal replacement therapy [14, 15]. Although we did not measure SBECD concentrations in this study, clinically significant accumulation of SBECD seems unlikely given the limited treatment duration and intermittent hemodialysis.

Overall, there were no signs of drug related toxicity. Markers for hepatic injury remained within the reference ranges during treatment and up to 8 days of follow up (Supplementary figure 2). An isolated increase in International Normalized Ratio (INR) was observed, which improved after vitamin K supplementation and therefore was not considered as treatment related (Supplementary figure 2). By day 5 of remdesivir treatment, oxygenation parameters had improved allowing for patient transfer to a hospital ward.

Our observation support the safety of remdesivir in patients with ESRD on hemodialysis, which will assist clinical decision making and guide future clinical trial design.

## Data Availability

Data will be made available upon reasonable request

