## Supplemental Material for "Pharmacokinetics of remdesivir in a COVID-19 patient with end-stage renal disease on intermittent hemodialysis"

### Supplementary table 1

General patient characteristics and medical history

| | |
|---|---|
| **Age** | 70-80 years |
| **Gender** | Male |
| **Weight** | 53.2 kg |
| **Glomerular Filtration Rate** | 0 mL/min (no residual renal function) |
| **Relevant medical history** | End-stage renal disease due to IgA-nephropathy<br>Intermittent hemodialysis for > 20 years<br>Paroxysmal atrial fibrillation<br>Type 2 diabetes mellitus<br>Coronary artery disease |

**Supplementary table 2**

Concomitant medication during remdesivir treatment

| Medication | Dose | Indication |
|---|---|---|
| Piperacillin/tazobactam | 2 x 4.5 g* | Suspected bacterial superinfection |
| Metoclopramide | 3 x 10 mg | Nausea |
| Dexamethasone | 1 x 6 mg | Severe COVID-19 |
| Famotidine | 1 x 20 mg | Stress ulcer prophylaxis |
| Acetylsalicyclic acid | 1 x 100 mg | Secondary prevention after NSTEMI |
| Levothyroxine | 1 x 75 µg | Hypothyroidism |
| Tilidin/naloxone | 2 x 100/8 mg | Chronic Pain Syndrome |
| Calcium acetate | 3 x 950 mg | Chronic kidney disease (phosphate binder) |
| Enoxaparin | 1 x 20 mg* | Thromboembolism prophylaxis |
| Heparine (during hemodialysis) | varying | Hemodialysis |

*Dose adjusted to GFR; NSTEMI, non-ST-elevation myocardial infarction

**Supplementary table 3**

Parameters of remdesivir (200 mg single dose) and its metabolites

| | Parameter | Index patient (no renal function) | Humeniuk et al. 2020[a] (healthy volunteers) |
|---|---|---|---|
| **Remdesivir (GS-5734)** | $AUC_{0-\infty}$* | 13.0 µg/mL*h | 3.3 µg/mL*h |
| | $C_{max}$ | 19.8 µg/mL | 7.8 µg/mL |
| | Clearance | 257 mL/min | 1000 mL/min |
| | Volume of distribution ($V_z$)* | 24.5 L | 87 L |
| | Apparent elimination half-life* | 1.10 h | 1 h |
| **GS-441524** | $AUC_{0-20.1\ h}$ | 18.4 µg/mL*h | n.a. |
| | $C_{max}$ | 1.15 µg/mL*h | 0.2 ng/mL |
| | Apparent elimination half-life | not estimable (∞) | 27 h |
| **GS-704277** | Apparent elimination half-life* | 2.98 h | 1.3 h |

[a] values reported for the 30 min infusion of 75 mg extrapolated to a 200 mg dose assuming dose linearity[1]; clearance was calculated from reported parameters as dose/AUC and volume of distribution as half-life * clearance /ln(2); * estimated using the last 3 time points with quantifiable remdesivir concentrations; $AUC_{0-\infty}$, area under the concentration vs. time profile extrapolated to infinity; $C_{max}$ , maximal observed plasma concentration; $V_z$, volume of distribution at pseudo-equilibrium

**Supplementary table 4**

Quantification of systemic remdesivir prodrug (GS-5734) and GS-441524 metabolite, relative quantification of remdesivir intermediate metabolite (GS-704277)

| Sample ID | Sampling time | Time after start infusion (h) | Remdesivir (ng/mL) | GS-441524 (ng/mL) | Ratio GS-704277/IS |
|---|---|---|---|---|---|
| 1 | day 1 - 13:02 | 0 | BQL | BQL | - |
| 2 | day 1 - 13:22 | 0.25 | 19845.3 | 46.208 | 0.955 |
| 3 | day 1 - 13:36 | 0.48 | 15513.7 | 145.65 | 3.028 |
| 4 | day 1 - 13:50 | 0.72 | 8525.0 | 264.93 | 4.392 |
| 5 | day 1 - 14:05 | 0.97 | 2918.6 | 375.16 | 4.234 |
| 6 | day 1 - 14:35 | 1.47 | 1484.9 | 460.43 | 3.227 |
| 7 | day 1 - 15:32 | 2.42 | 439.80 | 883.10 | 1.989 |
| 8 | day 1 - 17:32 | 4.42 | 127.69 | 958.27 | 1.613 |
| 9 | day 1 - 19:43 | 6.60 | 31.729 | 859.15 | 0.756 |
| 10 | day 2 - 07:06 | 17.98 | BQL | 1152.7 | 0.229 |
| 11 | day 2 - 09:14 | 20.12 | BQL | 876.44 | 0.148 |
| 12 | day 2 - 10:59 | 21.87 | BQL | 856.22 | - |
| 15 | day 2 - 13:55 | 24.80 | BQL | 589.44 | - |
| 18 | day 2 - 16:45 | 27.63 | BQL | 442.62 | - |
| 19 | day 2 - 17:03 | 27.93 | 8327.6 | 430.08 | 0.358 |
| 20 | day 2 - 17:15 | 28.13 | 6548.0 | 404.21 | 2.272 |
| 21 | day 2 - 17:29 | 28.37 | 3225.5 | 611.62 | 2.953 |
| 22 | day 3 - 16:00 | 50.88 | BQL | 1659.5 | - |
| 24 | day 4 - 16:30 | 75.38 | BQL | 1613.5 | - |
| 26 | day 5 - 09:55 | 91.80 | BQL | 1787.0 | - |
| 27 | day 5 - 11:34 | 93.45 | BQL | 1118.1 | - |
| 30 | day 5 - 13:40 | 95.55 | BQL | 835.26 | - |
| 33 | day 5 - 14:45 | 96.63 | BQL | 810.75 | - |
| 36 | day 5 - 15:05 | 96.97 | BQL | 549.99 | - |
| Dialysate | day 2 - 17:25 | - | BQL | 197.93 | - |

Abbreviations: BQL, Below Quantification Limit (remdesivir: 3.60 ng/mL, GS-441524: 10.8 ng/mL); IS, internal standard

**Supplementaty table 5**

Quantification of remdesivir prodrug (GS-5734) and GS-441524 metabolite in arterial (inlet) and venous (outlet) lines of the hemodialysis system

| Sample ID | Sampling time | Time after start infusion (h) | Remdesivir (ng/mL) | GS-441524 (ng/mL) |
|---|---|---|---|---|
| 13 HDA | day 2 - 11:05 | 21.97 | BQL | 792.30 |
| 14 HDV | day 2 - 11:05 | 21.97 | BQL | 334.94 |
| 16 HDA | day 2 - 13:55 | 24.80 | BQL | 540.66 |
| 17 HDV | day 2 - 13:55 | 24.80 | BQL | 229.09 |
| 28 HDA | day 5 - 11:34 | 93.45 | BQL | 1217.3 |
| 29 HDV | day 5 - 11:34 | 93.45 | BQL | 395.95 |
| 31 HDA | day 5 - 13:40 | 95.55 | BQL | 862.97 |
| 32 HDV | day 5 - 13:40 | 95.55 | BQL | 459.22 |
| 34 HDA | day 5 - 14:45 | 96.63 | BQL | 920.17 |
| 35 HDV | day 5 - 14:45 | 96.63 | BQL | 549.99 |

Abbreviations: HDA, hemodialysis arterial line (inlet); HDV, hemodialysis venous line (outlet); BQL, Below Quantification Limit (remdesivir: 3.60 ng/mL, GS-441524: 10.8 ng/mL)

**Supplementary figure 1**

Chest computed tomography (CT) performed eight days after diagnosis

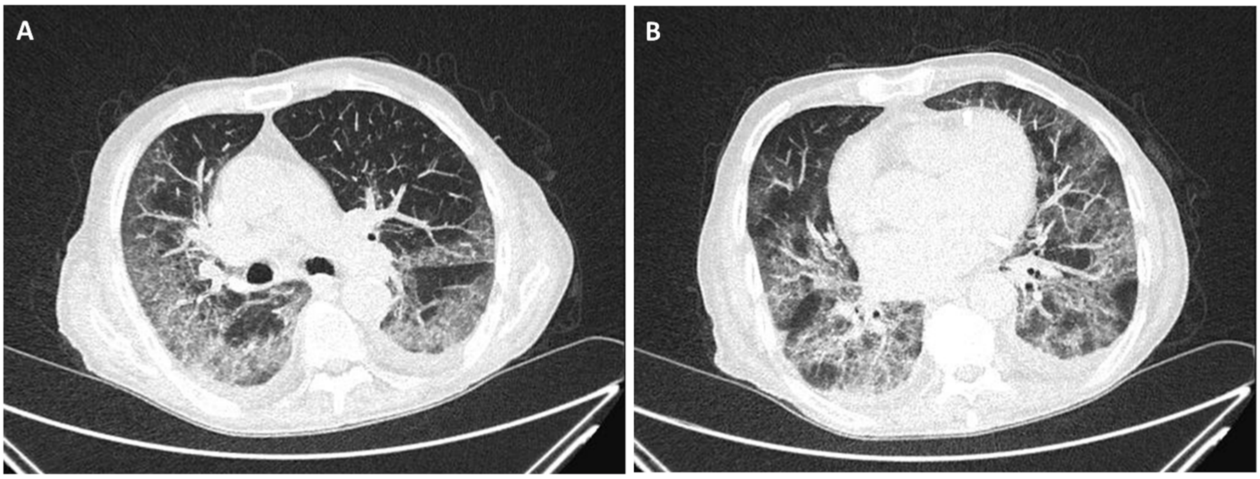

A (cranial) and B (caudal) pictures from low-dose plain CT scan performed eight days after hospital admission showing progressive bipulmonary ground glass opacities and consolidations with a predominantly peripheral distribution pattern. In addition, linear intralobular septal thickening (crazy paving) and small pleural effusions.

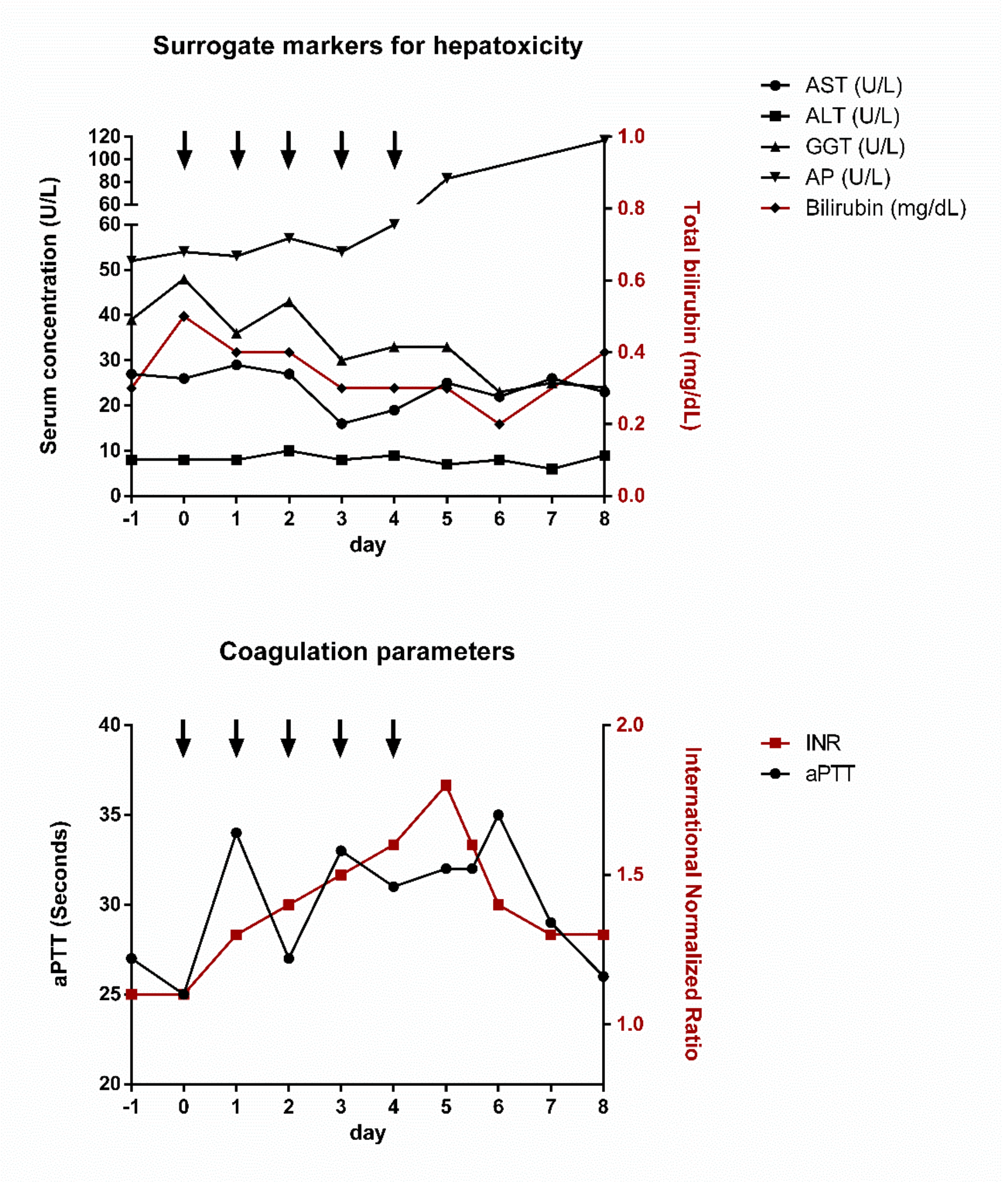

**Supplementary figure 2.** Daily monitoring of toxicity markers during antiviral treatment (day 0-4, ↓) of a patient with end-stage renal disease with the standard regimen of remdesivir (200 mg d0, 100 mg d1-4 IV) and follow up (day 5-8). Hemodialysis was performed on days 1 and 4. Laboratory reference ranges are not shown here but were only exceeded in case of INR which nearly normalized under vitamin K supplementation. Abbreviations: AST, aspartate transaminase; ALT, alanine aminotransaminase; GGT, gamma-glutamyltransferase; AP, alkaline phosphatase; Bilirubin = total bilirubin; INR, International Normalized Ratio, aPTT, activated partial thromboplastin time.
